# Genomic surveillance of Nevada patients revealed prevalence of unique SARS-CoV-2 variants bearing mutations in the RdRp gene

**DOI:** 10.1101/2020.08.21.20178863

**Authors:** Paul D. Hartley, Richard L. Tillett, David P. AuCoin, Joel R. Sevinsky, Yanji Xu, Andrew Gorzalski, Mark Pandori, Erin Buttery, Holly Hansen, Michael A. Picker, Cyprian C. Rossetto, Subhash C. Verma

## Abstract

Patients with signs of COVID-19 were tested with CDC approved diagnostic RT-PCR for SARS-CoV-2 using RNA extracted from nasopharyngeal/nasal swabs. In order to determine the variants of SARS-CoV-2 circulating in the state of Nevada, 200 patient specimens from COVID-19 patients were sequenced through our robust protocol for sequencing SARS-CoV-2 genomes. Our protocol enabled sequencing of SARS-CoV-2 genome directly from the specimens, with even very low viral loads, without the need of culture-based amplification. This allowed the identification of specific nucleotide variants including those coding for D614G and clades defining mutations. These sequences were further analyzed for determining SARS-CoV-2 variants circulating in the state of Nevada and their phylogenetic relationships with other variants present in the united states and the world during the same period of the outbreak. Our study reports the occurrence of a novel variant in the nsp12 (RNA dependent RNA Polymerase) protein at residue 323 (314aa of orf1b) to Phenylalanine (F) from Proline (P), present in the original isolate of SARS-CoV-2 (Wuhan-Hu-1). This 323F variant is found at a very high frequency (46% of the tested specimen) in Northern Nevada. Functional significance of this unique and highly prevalent variant of SARS-CoV-2 with RdRp mutation is currently under investigation but structural modeling showed this 323aa residue in the interface domain of RdRp, which is required for association with accessory proteins. In conclusion, we report the introduction of specific SARS-CoV-2 variants at a very high frequency within a distinct geographic location, which is important for clinical and public health perspectives in understanding the evolution of SARS-CoV-2 while in circulation.

## INTRODUCTION

Severe Acute Respiratory Syndrome coronavirus 2 (SARS-CoV-2), the cause of coronavirus disease 2019 (COVID-19), was first identified and reported in December 2019 in Wuhan, Hubei province, China [1-3]. RNA sequencing and phylogenetic analysis of specimens taken during the initial outbreak in Wuhan determined that the virus is most closely related (89.1% nucleotide similarity) to a group of SARS-like coronaviruses (genus Betacoribavirus, subgenus Sarbecovirus) which had previously been identify in bats in China [4,2]. Coronaviruses have a recent history as emerging infections, first SARS-CoV in 2002-2003, and Middle East respiratory syndrome coronavirus (MERS-CoV) in 2012, both zoonotic infections that cause severe respiratory illness in humans [5,6,2,7,8] [2,5-8]. Unlike SARS-CoV and MERS-CoV which displayed limited global spread, SARS-CoV-2 has spread around the world within a few months. There are specific characteristics of SARS-CoV-2 which have facilitated the transmission, including infections that result in asymptomatic or mild disease, allowing for under-characterized transmission.

SARS-CoV-2 is an enveloped, positive single-stranded RNA virus. Detection of SARS-CoV-2 in patients has primarily occurred using RT-qPCR to detect viral RNA from respiratory specimens (primarily nasal and nasopharyngeal swabs). While RT-PCR results can be quantified through determination of a cycle threshold (Ct) value for each sample, it does not yield sequence data leading to the description of genomic variants. To further study of such variants, and to better understand the epidemiology of the virus in the state of Nevada, we developed a workflow that allowed us to sequence SARS-CoV-2 genomic RNA from patient swabs containing a broad range of viral loads. Of the sequences of SARS-CoV-2 currently submitted to common database (GenBank and GISAID), several were obtained after the virus had been passed in Vero cells [9,10] and others came directly from patient specimens [11,12]. Certain data have suggested a potential for lab acquired mutations following passage in cell culture [13,9]. Specifically, a report of SARS-CoV-2 passage in Vero cells which resulted in a spontaneous 9 amino acid deletion within the spike (S) protein that overlaps with the furin cleavage site [13]. The loss of this site is suggested to increase the viral entry into Vero cells [14]. For both research and epidemiological purposes, sequencing of SARS-CoV-2 directly from patient specimens not only reduces the possibility of laboratory acquired mutations following passage in cell culture but also reduces the time that would be spent growing the virus from the patient specimens and subsequently also reduces handling larger amounts of infectious virus. Additionally, one of the goals in developing an optimized SARS-CoV-2 NGS protocol was to be able to generate adequate depth of coverage of the viral genome while minimizing the sequencing of non-viral RNA which would allow for more specimens to be multiplexed together during sequencing.

Our workflow employs a combination of RNA amplification, conversion into Illumina-compatible sequencing libraries and enrichment of SARS-CoV-2 library molecules prior to sequencing. Using this novel methodology, we sequenced SARS-CoV-2 from a total of 200 patient specimens collected over a three-month period originating from Nevada. Of the 200 selected, 173 were sequenced with enough quality to be used for determining SARS-CoV-2 nucleotide variants to perform further phylogenetic analysis and study the viral epidemiology within the state of Nevada. Analysis of the data suggests a specific epidemiological course for the local epidemic within Northern Nevada. This was characterized by an initial observation of variants closely resembling isolates originating directly from China or Europe. Subsequent to government-mandated period of restrictions on business and social activity, we observed that a viral isolate not seen elsewhere in the world emerged within Northern Nevada cases (nucleotide 14,407 and 14,408). This isolate contains an amino acid change in residue P323L/F of RdRp (nsp12). Furthermore, we found that sampled viral isolates in Southern Nevada, unlike those in Northern Nevada, closely resembled the makeup of the United States in general.

## MATERIALS AND METHODS

### SARS-CoV-2 specimen and library preparation

Nasal and Nasopharyngeal swab specimens were received at the Nevada State Public Health Lab (NSPHL) or Southern Nevada Public Health Lab (SNPHL) and RNA extraction was completed using either a QIAamp Viral RNA Mini Kit (QIAGEN) or Mag-Bind Viral DNA/RNA kit (Omega Biotek). Specimens were tested for the presence of coronaviral RNA using FDA-approved kits that employed RT-PCR to detect SARS-COV-2 RNA.

A set of 200 coronavirus positive specimens were selected for genome sequencing. Specimens were treated with DNase I (QIAGEN) for 30 minutes at room temperature and concentrated using RNeasy Minelute spin columns (QIAGEN) based on the manufacturer supplied protocol. These concentrated samples were converted into Illumina-compatible sequencing libraries with a QIAseq FX Single Cell RNA Library kit (QIAGEN). RNA samples were annealed to a 1:12.5 dilution of QIAseq FastSelect -HMR probes (QIAGEN) to reduce subsequent amplification of human ribosomal RNA. After treatment to remove trace DNA from the samples, a reverse transcription reaction was carried out using random hexamers. The synthesized DNA was ligated to one another, followed by isothermal linear amplification. Amplified DNA (1 μg) was enzymatically sheared to an average insert size of 300 bp, and Illumina-compatible dual-indexed sequencing adapters were ligated to the ends. Next, about 300 ng of adapter-ligated sample was amplified with 6 cycles of PCR with KAPA HiFi HotStart polymerase (Roche Sequencing Solutions). Enrichment of library molecules containing SARS-CoV-2 sequence was conducted with a myBaits kit and coronavirus-specific biotinylated probes (Arbor Biosciences). Each enrichment used 500 ng of PCR-amplified DNA, was carried out based on manufacturer instructions at a hybridization temperature of 65° for 16 hours, and was completed with 8-16 cycles of PCR using KAPA HiFi HotStart polymerase. Samples were sequenced using an Illumina Next-seq mid-output (2 × 75). The generated FASTQ files from the sequencing reaction were analyzed as described below. The data files are available at GISAID, NCBI under the https://www.ncbi.nlm.nih.gov/bioproject/657893

### Computational analysis

Sequence pair libraries were trimmed using Trimmomatic, version 0.39 and adapter-clipping setting “2:30:10:2:keepBothReads” [15]. Read pairs were aligned against the Wuhan reference genome (NC_045512.2) by Bowtie 2, version 2.3.5, local alignment [16]. PCR optical duplicates were removed via Picard MarkDuplicates [17].

Variants were called using Freebayes, version 1.0.2, with ploidy set to 1, minimum allele frequency 0.75, and minimum depth of 4 [18]. No variants were called in the first 200 bp and final 63 bp of the COVID-19 genome. High-quality variant sites were selected where site “QUAL > 20” using *vcffilter*, VCFlib version 1.0.0_rc2 [19]. Individual genomes were reconstructed by their filter-passing variants using *bcftools consensus* and only where aligned coverage depth ≥ 4; bases with coverage below four are reported as unknown (Ns) [20].

A set of 3,644 complete, high-coverage SARS-CoV-2 genomes reported in the July 15, 2020 Nextstrain.org global subsample and metadata were obtained from GISAID and combined with our own samples to determine their phylogenetic placement [21,22]. Four of the global samples were set aside after screening for unexpected FASTA characters. The combined sets of global and Nevadan samples were aligned together, with metadata, by the *augur* phylodynamic pipelines of the *ncov* build of the *nextstrain* command-line tool, version 2.0.0.post1 [23].

Cumulative frequency of D614G, clades (19A, 19B, 20A, 20B, 20C), and P323L/F were calculated at each time point based on the total number of specimens up to the indicated date. Plots and pie charts were generated using GraphPad Prism (version 8).

### nsp12 protein modeling

Sequence of nsp12 (RdRp) protein for SARS-CoV-2 (YP_009725307.0) was retrieved from NCBI protein database and 3D model was structured based on a previously published report (PDB ID: 6XEZ [24]. In addition to nsp12 (chain A), the model also contains nsp7 (chain C), nsp8 (chain B and D), nsp13 (chain E and F), ligands (Zn^2+^, Mg^2+^) and RNA template and product strands. Mutational changes to residue 323 within nsp12 were performed using PyMol Molecular Graphics System (version 2.0, Schrödinger LLC). The original proline (P) was mutated to either leucine (L) or phenylalanine (F) as indicated, these residues along with residues containing side chains within 5 Å of P323L/F are shown as sticks. The rotamers for each P323L/F were assessed and those with the least rotational strain and steric hindrance were used to generate the final image. To determine any NCBI deposited sequences which contain the P323F variant, standard protein BLAST from the BLASTp suite was used to find nsp12 protein sequences which contained FSTVFPFTSFGP (P323F is bold and underlined) from full length SARS-CoV-2 genomes. The P323F amino acid changes were confirmed with the NCBI deposited nucleotide sequences.

## RESULTS

### RNA-seq workflow and assembly of SARS-CoV-2 genomes

A total of 200 SARS-CoV-2 positive specimens collected in Nevada from March 6 to June 5 were randomly selected to have their viral genomes sequenced for variant analysis and subsequent epidemiological studies (Fig 1A and Materials & Methods). Of the sequenced specimens, 173 had >90% coverage and sufficient depth to accurately call those genomic positions with variants (Fig. 1B). An alignment of SARS-CoV-2 genomes from these specimens is presented as supplemental data (Supplemental Fig. S1) showing 173 specimens with over 90% coverage. These 173 specimens represented 133 patient specimens from Northern Nevada (including Washoe County of which Reno is the major city, the Carson-Tahoe area, and other northern, rural counties), 40 patient specimens from Southern Nevada (Clark County, which encompasses Las Vegas and surrounding cities). Nucleotide similarity and variants were determined and used to measure the phylogenetic relationships (Supplemental Fig. S2). The combined nucleotide diversity across the entire SARS-CoV-2 genome for the Nevada specimens is shown in figure 1D, along with the genomic areas that were assessed for change in frequency corresponding to amino acids D614G, P323L/F and nucleotide 379.

**Figure 1.**
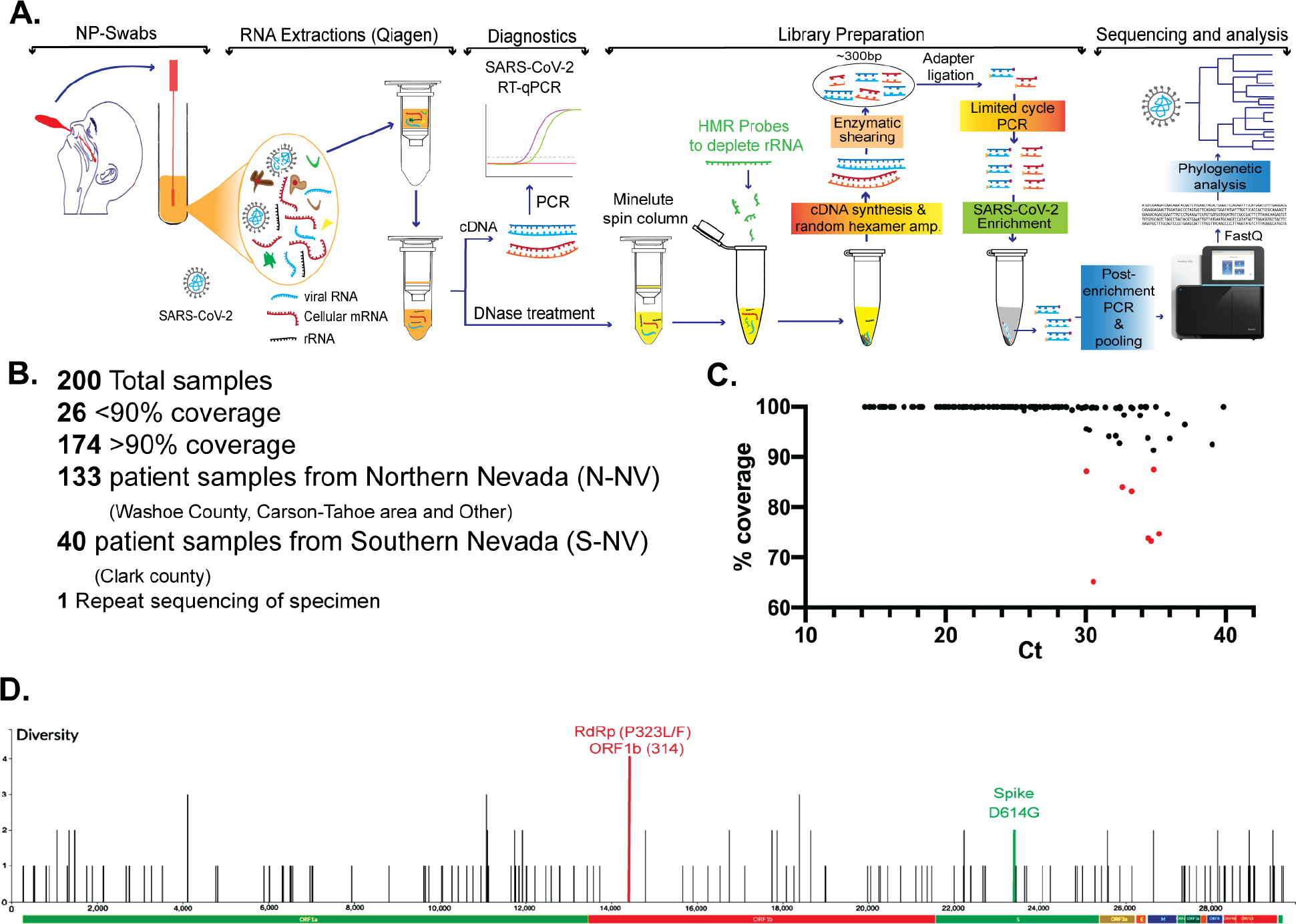
Workflow of SARS-CoV-2 genome sequencing and analysis from nasopharyngeal patient specimens in Nevada. (A) RNA was extracted from Nasal or Nasopharygeal (NP) swabs taken from patients in Nevada and first used to determine the presence of SARS-CoV-2 genomes by RT-qPCR. Next generation sequencing (NGS) libraries were prepared from positive specimens, this included steps for ribosomal RNA depletion and SARS-CoV-2 enrichment. Subsequent libraries were pooled and used for whole genome sequencing at the Nevada Genomics Center on the Illumina NextSeq 500 instrument. FASTQ files were aligned to the reference genome, and analyzed to determine nucleotide variation and phylogenetic relationship. (B) A total of 200 specimens were sequenced, of which 174 had over 99% coverage of the SARS-CoV-2 genome. This included 133 patient specimens from Northern Nevada, 40 from Southern Nevada and 1 specimen that was re-sequenced. (C) Correlation between RT-qPCR Ct value and the percentage of coverage in the whole genome sequencing after trimming and alignment. (D) Nucleotide variants across the SARS-CoV-2 genome in the 173 specimens from Nevada from March 6 to June 5.

During the sequencing analysis we also examined the correlation between Ct values from the diagnostic RT-PCR and percentage coverage of the viral genome to determine the performance and robustness of our sequencing method in relation to available viral RNA in a specimen of a given Ct (Fig. 1C). On average, a Ct value less than 37 resulted in at least 90% coverage to the SARS-CoV-2 genome. Importantly, our in-house developed method for viral genome enrichment and sequencing directly from the patient’s specimens (nasal and nasopharyngeal swabs) was robust and yielded sequences covering over 90% of the genome even in samples having very high Ct (∼40) of viral genome detection. This is highly significant and shows the power of our workflow in sequencing of SARS-CoV-2 genome from a spectrum of samples including the ones having inadequate amounts of specimen (due to the variability in collection) or lower viral loads in nasal secretions. Consequently, our sequencing protocol avoids any molecular epidemiological bias, which may get acquired through cell culture-based amplification especially in those specimens with high Ct (low viral load) as our method eliminates the need of virus culture.

### Prevalence of amino acid variant D614G of SARS-CoV-2 spike protein in specimens collected in Nevada

Earlier studies have revealed the emergence, spread and potential importance of an alteration, D614G (genomic change at 23403A>G), of the spike protein [25]. This missense mutation has become a clade-distinguishing locus that differentiates viral isolates originating in Asia from those that have emerged from Europe. A total of 173 cases were analyzed to determine the number and relative proportion of the specimens which carried the D614G spike protein variant in Nevada. The cumulative frequency for D614 and G614 were plotted from March 6 to June 5 (Fig. 2A). Specimens from the beginning of March represent the earliest known cases in Nevada, and of the 14 specimens sequenced during this time period (March 6-March 15) D614 was the predominant variant. This shifted from March to June with an increasing frequency of the G614 allele. The trend for specimens originating from either Northern Nevada (N-NV) and Southern Nevada (S-NV) both show a higher frequency of G614 (Fig. 2B). We used a subsampling of sequence data from Nextstrain.org to assess the frequency of D614G in the United States and globally during the same time period (March 6 to June 5) (Fig. 2B and 2C). The global trend by continent of D614G is also similar, with G614 at a higher frequency, the one noted exception is in Asia, where D614 and G614 continue to exist in equal proportions (Fig. 2C).

**Figure 2.**
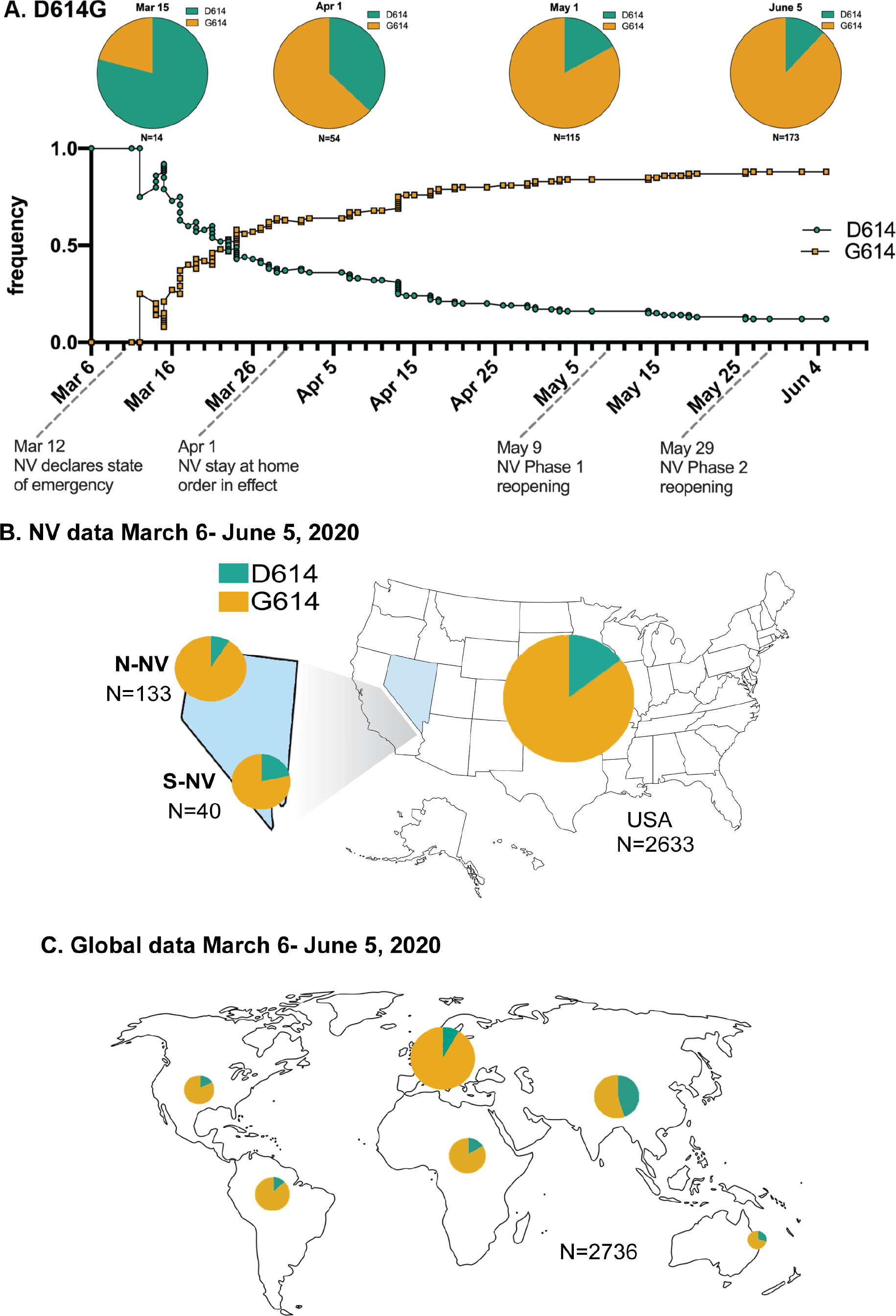
Distribution of D614G in Nevada and comparison with the United States and global proportion. (A) Cumulative frequency of D614G in 173 patient specimens from Nevada from March 6 to June 5, 2020 (D614 is indicated by teal, G614 is indicated by yellow). Pie charts depict the cumulative proportion up to the indicated time point (March 15, April 1, May 1, June 5). The total number of specimens included at each time point is specified below each pie chart. Effective dates of emergency orders and regulatory responses to SARS-CoV-2 spread in Nevada are indicated on the frequency graph time axis. (B) Proportion of D614G in the United States from March 6 to June 5, specimens from Nevada are divided in the geographic area that they originated from, Northern Nevada (N-NV) includes 133 specimens from Washoe County, Carson-Tahoe, and other northern counties, and Southern Nevada (S-NV) includes 40 specimens from Clark County. (C) Global proportion of D614G in the shown regions during the same time period from a subsampling of sequences deposited in Nextstrain.org. The size of the pie chart corresponds to the relative specimen number for each region.

### Frequency of SARS-CoV-2 clades in Nevada

Worldwide, there are currently 5 main clades (19A, 19B, 20A, 20B, 20C) of SARS-CoV-2 differentiated based on specific nucleotide profiles in the Year-letter scheme of https://clades.nextstrain.org. Clade 19A and 19B are defined by C8782T and T28144C, respectively. 20A is a derivative of 19A and contains mutations C3037T, C14408T and A23403G (resulting in D614G). 20B is defined by mutations G28881A, G28882A and G28883C, and 20C contains C1059T and G25563T [26].

To assess the introduction and spread of the clades in Nevada the cumulative frequency for the clades were plotted from March 6 to June 5 (Fig. 3A). The earliest sequenced specimens from Nevada were collected in the beginning of March (March 6-March 15) and are predominantly from clades 19A and 19B. Additional sequenced specimens collected from March to June revealed a shift to a higher frequency of 20C (Fig. 3A). We performed phylogenetic reconstruction of the Nevada specimens and differentiated the clades on the circular dendrogram by color (Fig. 3B). There were discordant trends in the dominant clade for specimens originating from either Northern Nevada (N-NV) and Southern Nevada (S-NV) (Fig. 3C and 3D). Specimens from Northern Nevada (Washoe County, Carson-Tahoe, and other counties) showed a prevalence of 20C, while the Southern Nevada specimens from Clark County had a larger proportion of 20A (Fig. 3C and Supplemental Fig. S3a). We used a subsampling of Nextstrain.org data to assess the frequency of clades in the United States and globally by continent during the same time period (March 6 to June 5). The dominant clade in the United States was 20C, similar to the frequency seen in the total Nevada samples (N-NV and S-NV) (Fig. 3A and 3C). The global clade distributions were variable in areas outside of Asia while clades 19A and 19B are noted to be more prevalent in Asia. (Fig. 3D).

**Figure 3.**
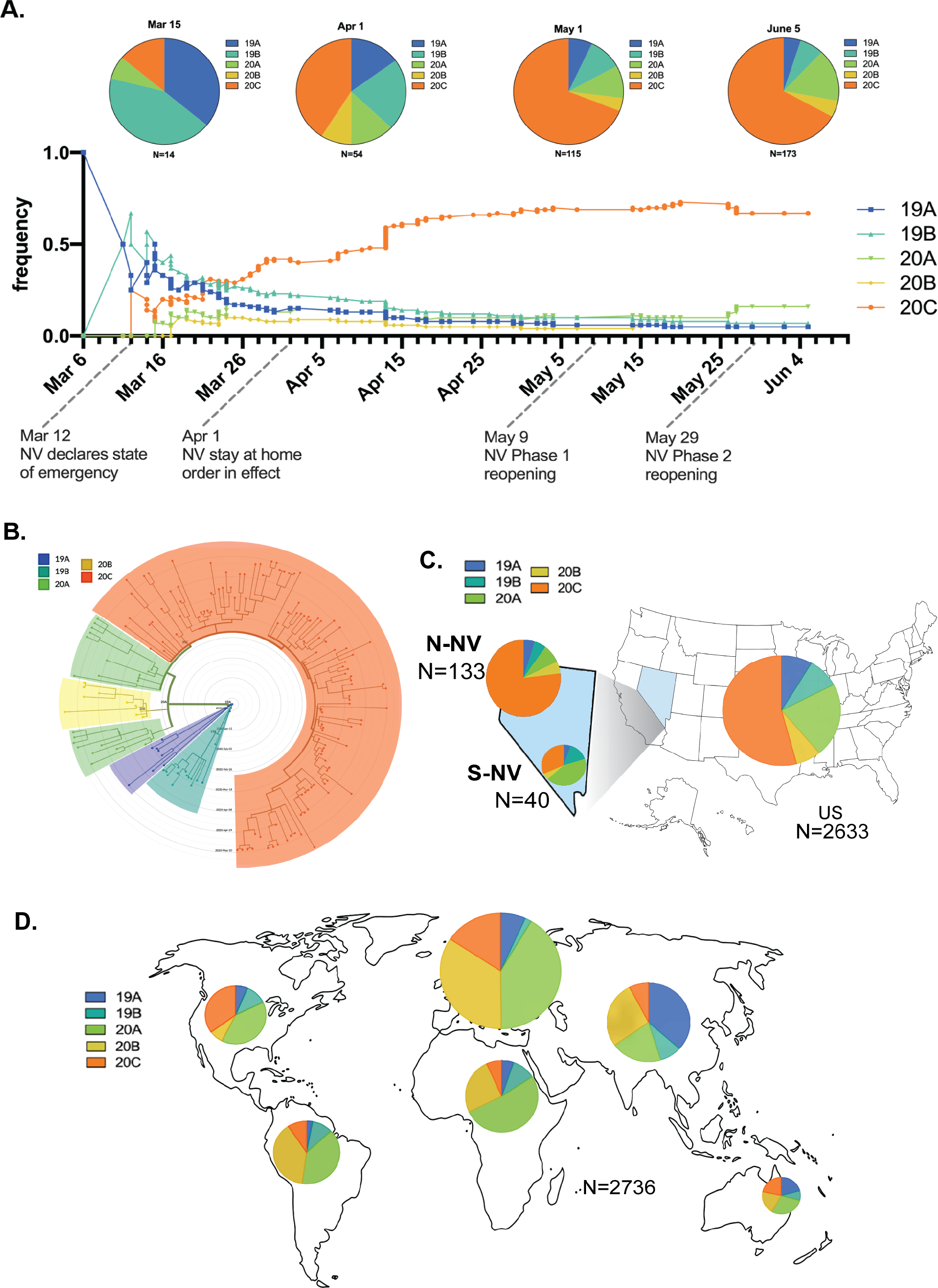
Distribution of SARS-CoV-2 clades in Nevada. (A) Cumulative frequency of SARS-CoV-2 clades in 173 patient specimens from Nevada during March 6 to June 5. The five clades are colored 19A (blue), 19B (teal), 20A (green), 20B (yellow) and 20C (orange). Pie charts depict the cumulative proportion up to the indicated time point (March 15, April 1, May 1, June 5). The total number of specimens included at each time point is specified below each pie chart. Dates of emergency orders and regulations meant to slow the spread of SARS-CoV-2 in Nevada are indicated on the time scale of the frequency graph. (B) Circular dendrogram depicting clades from Nevada specimens. (C) Pie chart of the clades from northern Nevada (N-NV), southern Nevada (S-NV) and the United States. (D) Pie charts show the proportion of clades from global regions during the same time period from a subsampling of sequences deposited in Nextstrain.org. The size of the pie chart corresponds to the relative specimen number for each region.

### Prevalence of amino acid variant P323L/F of SARS-CoV-2 nsp12 (RdRp) in Nevada

Analysis of sequencing data revealed a novel observation for our specimens at bases 14,407 and 14,408 which results in a change at residue 323 in nsp12 (RdRp). For the Wuhan isolate at 14,407 and 14,408 there is CC for proline (P), the variants have CT for leucine (P323L) and TT for phenylalanine (P323F). To assess the introduction and spread of P323L/F in Nevada, the cumulative frequency of P323, L323 and F323 were plotted from March 6 to June 5 (Fig. 3A). Nevada specimens from the beginning of March (March 6-March 15) showed P323 to be the predominant variant. As additional specimens were collected and sequenced from March to June there was a shift to a higher frequency of L323 and F323 (Fig. 4A). We performed phylogenetic reconstruction of the Nevada specimens and noted the P323L/F variants on the circular dendrogram with the indicated colors (Fig. 4B). Interestingly, analysis of the Northern Nevada and Southern Nevada specimen showed very different dominant variants (Fig. 4B). In Northern Nevada the F323 was more prevalent, while in Southern Nevada L323 was more prevalent. We used a subsampling of Nextstrain.org data to assess the frequency of P323L/F in the United States and globally during the same time period (March 6 to June 5). P323 was the predominant variant in Asia, while L323 was more prevalent in other areas of the world and F323 was only appreciably noted in North America (Fig. 4D and 4E).

**Figure 4.**
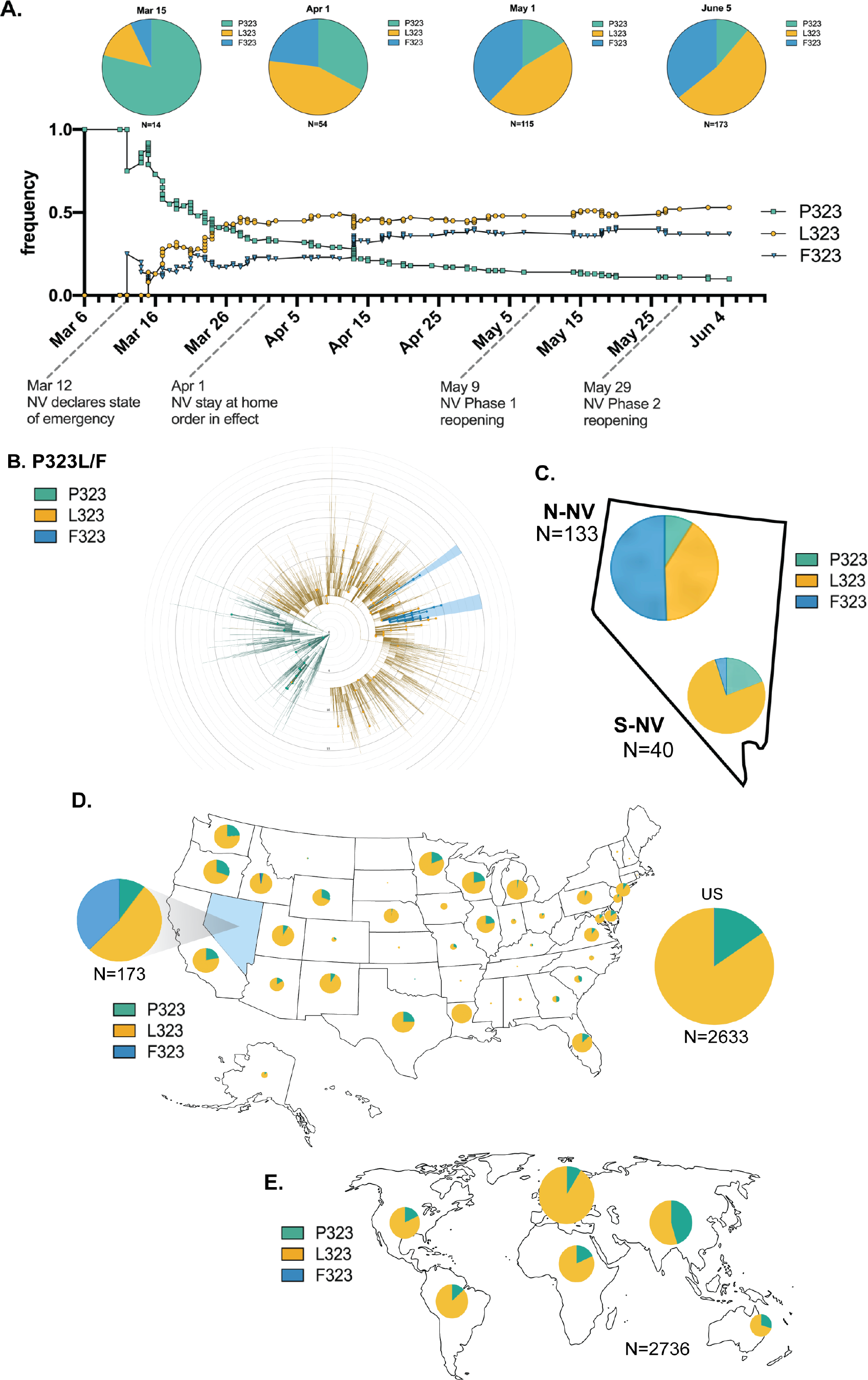
Distribution of P323L/F (nsp12, RdRp) in Nevada. (A) Cumulative frequency of P323L/F (nsp12, RdRp) in 173 patient specimens from Nevada during March 6 to June 5. The amino acid at position 323 is indicated by teal for proline (P), yellow for leucine (L) and blue for phenylalanine (F). Pie charts depict the cumulative proportion up to the indicated time point (March 15, April 1, May 1, June 5). The total number of specimens included at each time point is specified below each pie chart. Dates of emergency orders and regulations meant to slow the spread of SARS-CoV-2 in Nevada are indicated on the time scale of the frequency graph. (B) Circular dendrogram representing the distribution of amino acid change at residue 323 of nsp12 from a global subsampling of sequences deposited in Nextstrain.org from March 6 to June 5, the larger dots indicate specimens from Nevada. (C) Pie chart indicating the ratio of P/L/F in Northern NV and Southern NV specimens from this study. (D) Proportion of P323L/F from a subsampling of sequences deposited in Nextstrain.org for the United States and (E) global regions from March 6 to June 5. The size of the pie chart corresponds to the relative specimen number for each region.

While we are investigating the phenotypic significance of this predominant variant of RdRp, we performed in-silico structural modeling of RdRp to determine the spatio-temporal location of this 323aa on RdRp in complex with its accessory proteins, nsp7, nsp8 and nsp3. Our data showed the location of 323aa in the interface domain of RdRp and variation of P323 to L or F did not significantly change the conformation of the protein (Fig. 5). Since the interface domain (aa 251-398) acts as a protein-interaction junction for the finger domain of the polymerase and the second subunit of nsp8 (accessory protein), required for the polymerase activity, we anticipate this mutation to have phenotypic effect on the RdRp activity. This suggested that P323 variants of RdRp may have altered phenotype with fitness advantage/disadvantage in transmission or pathogenicity and pending investigation will provide confirmatory results on this highly prevalent mutation.

**Figure 5.**
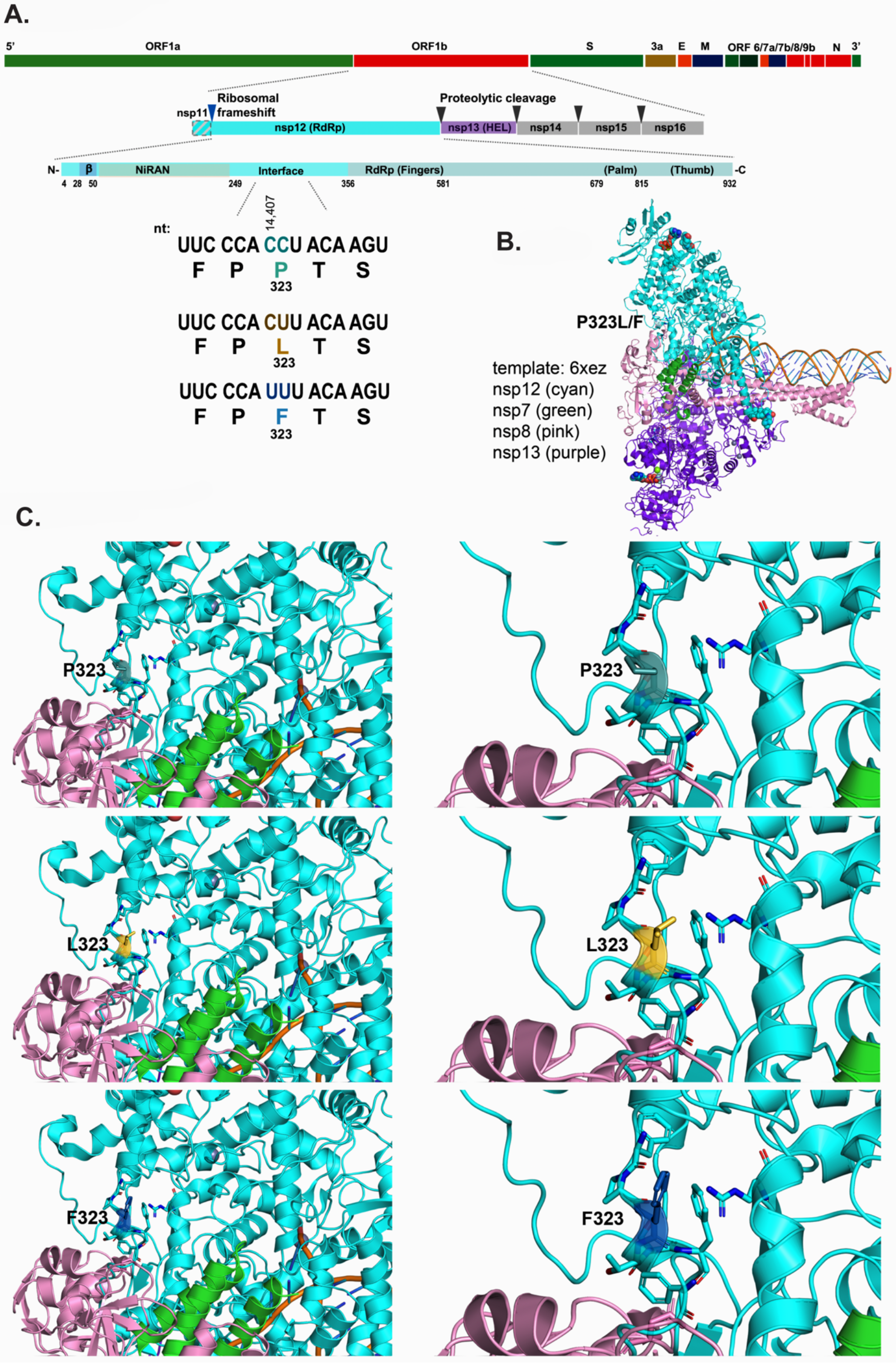
Structure of SARS-CoV-2 nsp12 (RdRp) P323L/F. (a) Diagram depicting ORF1b genomic location and encoded proteins. Below the linear protein schematic of nsp12 specific nucleotide variants at position 14,407 and 14,408 and the resulting amino acid changes are indicated. (b) SARS-CoV-2 replicase complex modeled from 6XEZ template. This model includes nsp12, nsp7, nsp8, nsp13, ligands (ZN, Mg), the template and product strand of RNA. P323L/F within the interface domain of nsp12 is located at the left side of the complex. (c) Model nsp12 showing P323L/F within the interface domain. Residue 323 is shown with either P, L or F and amino acids with side chains within 5 Å of residue 323 are depicted as sticks in the cartoon model.

## DISCUSSION

We have developed a novel method, which combines specific depletion and enrichment strategies that results in efficient SARS-CoV-2 RNA-seq with high genome coverage and depth. An advantage of this protocol is that it generates sequence data directly from swab specimens without the need to passage the virus in cell culture thereby reducing the handling of infectious material and induction of culture-acquired mutations. Another obstacle in sequencing directly from swab specimens is that most FDA-approved commercially available RNA extraction kits are specifically optimized to recover low amounts of total nucleic acids, include carrier polyA RNA that could be convertible into sequence able molecules, as has been observed previously with RNA-seq of Lassa- or Ebola-positive clinical specimens [27].

The workflow incorporates amplification of low-abundance RNA into micrograms of DNA, followed by conversion from a fraction of the DNA into Illumina-compatible sequencing libraries and enrichment of these libraries for SARS-CoV-2 sequences. In addition, during the reverse transcription step a reagent was incorporated to reduce the subsequent amplification of host ribosomal RNA. This approach is robust in that it converts low amounts of RNA into microgram quantities of DNA representative of all the RNA species (aside of rRNA) present in the specimen. This DNA can be stored indefinitely to be interrogated by multiple techniques at a later date. Additionally, RNA amplification is likely less sensitive to low viral abundance compared to RT-PCR. Finally, the use of probes to enrich for coronavirus-specific sequencing library molecules is less sensitive to variants compared to tiling PCR amplicon approaches [28-32].

The data herein implicate that early in the pandemic, before the “stay-at-home” order on April 1st, there were multiple introductions of SARS-CoV-2 into the state of Nevada. From April 1st to the beginning of June, Nevada experienced a period of semi-isolation, as the casinos and most hotels shut down, tourism and travel to the state essentially stopped. Because of the stay-at-home order and social distancing measures put in place, there was less mobility of people within and between states. It is possible that these measures, compounded by potential inherent transmission variability of some viral isolates, influenced the change in the frequency of D614G, clades and P323L/F that we noted during this time period within Nevada. In addition, we also found 379C>A with a high prevalence in our study specimens compared to the subsampling of sequences from the United States and globally (Figure 6). This is a synonymous mutation in nsp1, hence the biological relevance of this nucleotide variant remains to be elucidated.

**Figure 6.**
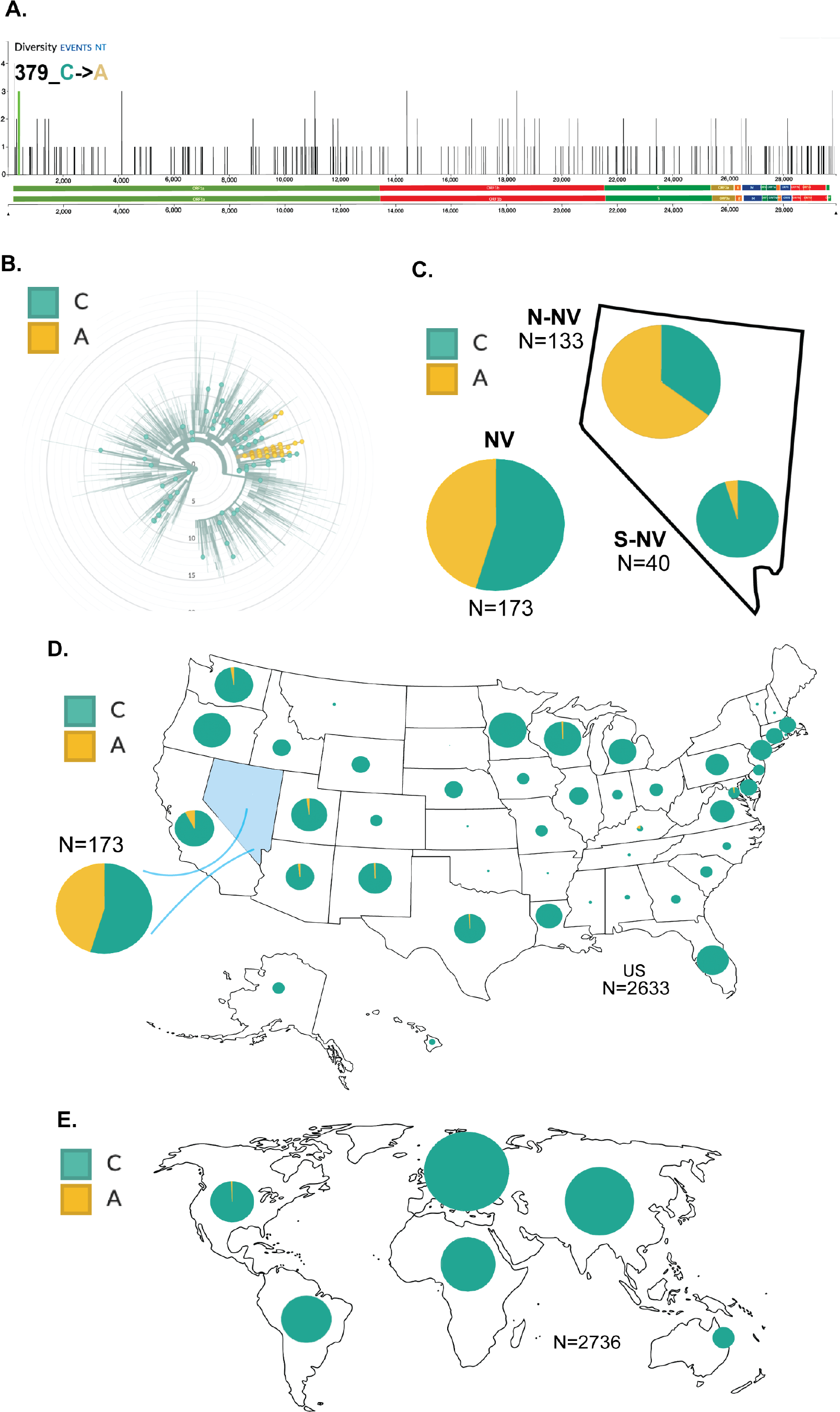
Distribution of nucleotide variant 379C>A. (a) Green line at the far-left end of the genome denoted nucleotide position 379 of nsp1. (b) Circular dendrogram of global subsample of sequences from Nextstrain.org with NV specimens indicated by larger dots. (c) Pie chart indicating the proportion of sequences with either the cytidine (C) or adenosine (A) at position 379 from the Nevada specimens. Subsample of sequences from Nextstrain.org were used to generate the proportion of 379C>A in (d) the indicated states within the U.S. and (e) internationally. The size of the pie chart corresponds to the relative specimen number for each region.

We found the overall trend of D614G in Nevada during this time period to be similar with what was observed in other states and internationally, with the exception of within Asia where the D614 allele had originated. We noted that there were differences between Northern Nevada and Southern Nevada. In Northern Nevada clade 20C and F323 were more frequent, while during this same time period in Southern Nevada clade 20A and L323 were more prevalent. These data indicate that there were distinct genomic profiles of the SARS-CoV-2 viruses that were circulating in these populations during the initial months of the pandemic while stay-at-home order were in place to help prevent transmission of the virus.

Of the 14,885 complete SARS-CoV-2 genomes available (as of August 14, 2020) in NCBI there are only 6 genomes that have the P323F variant (accession number: MT706208, LR860619, MT345877, MT627429, MT810889, MT811171). In this study 62 of the 133 specimens from Northern Nevada contain P323F. That is 46% of specimens from Northern Nevada contained P323F compared to 0.04% of NCBI deposited SARS-CoV-2 isolates. This was a significant accumulation of one specific SARS-CoV-2 variant in Northern Nevada, which could have been because of the circulation of this unique variant in the community without the introduction of new variants restricted by the shelter in place orders. However, regardless of the confined spread, P323F variant may have altered phenotypic characteristics, which have contributed to its increased prevalence and thus an active area of investigation. In an attempt to understand the role of this amino acid, our structural modeling of RdRp showed that 323aa is located in the interface domain, which acts as the junction for the interaction of accessory protein (nsp8), required for the polymerase activity [33]. Importantly, P323 of the Wuhan SARS-CoV-2 (Wuhan-Hu1) have mutated to Leucine (P323L) in all D614 variant of spike glycoprotein, which supposedly have higher transmission [25]. Although the role of D61G in combination with P323L of RdRp on viral transmission has not been investigated but co-existence of these mutational changes (D614G and P323L) in almost all predominantly detected variants of SARS-CoV-2 reflect their importance in transmission and pathogenicity. Consequently, higher prevalence of the P323 mutated to 323F (P323F) in variants circulating in the patients of Northern Nevada may suggest the importance of this specific amino acid in virus replication or transmission.

## Data Availability

https://www.ncbi.nlm.nih.gov/bioproject/657893

## DATA AVAILABILITY

All sequences are available at bio project: https://www.ncbi.nlm.nih.gov/bioproject/657893. All reported data are deposited and available at GISAID: hCoV-19/USA/NV-NSPHL-A (0004-0210)/2020.

## ACKNOWLEDGEMENTS

We thank the staff at the Nevada State Public Health Lab (NSPHL) and Southern Nevada, Public Health Laboratory (SNPHL) for providing the RNA extracts from the patient’s specimen. This work was supported by University of Nevada, Reno Vice President for Research and Innovation (VPRI), Department of Microbiology & Immunology, UNR School of Medicine, Nevada IDeA Network of Biomedical Research Excellence (INBRE) from the National Institute of General Medical Sciences (GM 103440 and GM 104944) from the National Institutes of Health (NIH). The authors wish to acknowledge the support of Research & Innovation and the Office of Information Technology at the University of Nevada, Reno for computing time on the Pronghorn High-Performance Computing Cluster.

## AUTHOR CONTRIBUTION

PDH: conceptualization, formal analysis, methodology, writing – original draft preparation, writing – review and editing

RLT: conceptualization, formal analysis, methodology, writing – original draft preparation, writing – review and editing

XY: formal analysis

DPA: conceptualization, funding

JRS: formal analysis, review and editing

AG: methodology

EB: specimen procurement and diagnostic testing

HH: specimen procurement and diagnostic testing

MP: conceptualization, specimen procurement, diagnostic testing, formal analysis, project administration, writing – review and editing

CCR: conceptualization, formal analysis, methodology, project administration, funding, writing – original draft preparation, writing – review and editing

SCV: conceptualization, formal analysis, methodology, project administration, funding, writing – review and editing

## DECLARATIONS

### Ethics Approval

Deidentified human specimens (nasal and nasopharyngeal swabs) were used for the extraction of viral RNA all the experiments were done in accordance with guidelines of the University of Nevada, Reno. The University of Nevada, Reno Institutional Review Board (IRB) reviewed this project and determined this study to be EXEMPT FROM IRB REVIEW according to federal regulations and University policy.

The Environmental and Biological Safety committee of the University of Nevada, Reno, approved methods and techniques used in this study.

## SUPPLEMENTARY FIGURES

### SUPPLEMENTARY INFORMATION

**Figure S1.**
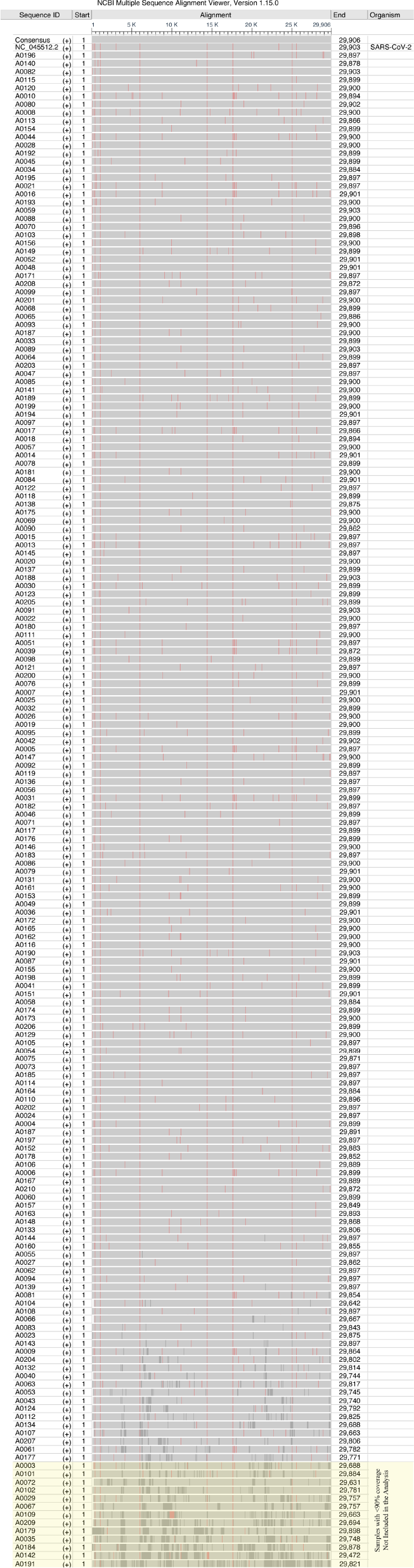
Alignment of SARS-CoV-2 sequences from NV patient’s specimens. SARS-CoV-2 sequences from patient specimen were aligned together with the original COVID-19 sequence from Wuhan, NC_045512.2, using multiple sequences alignment tool MUSCLE 3.8.31. The aligned sequences were sorted based on number of Ns in each sequence from the smallest to the largest. The sorted alignment file in aln format was uploaded to the web portal of NCBI Multiple Sequence Alignment Viewer, Version 1.15.0, for visualization.

**Figure S2.**
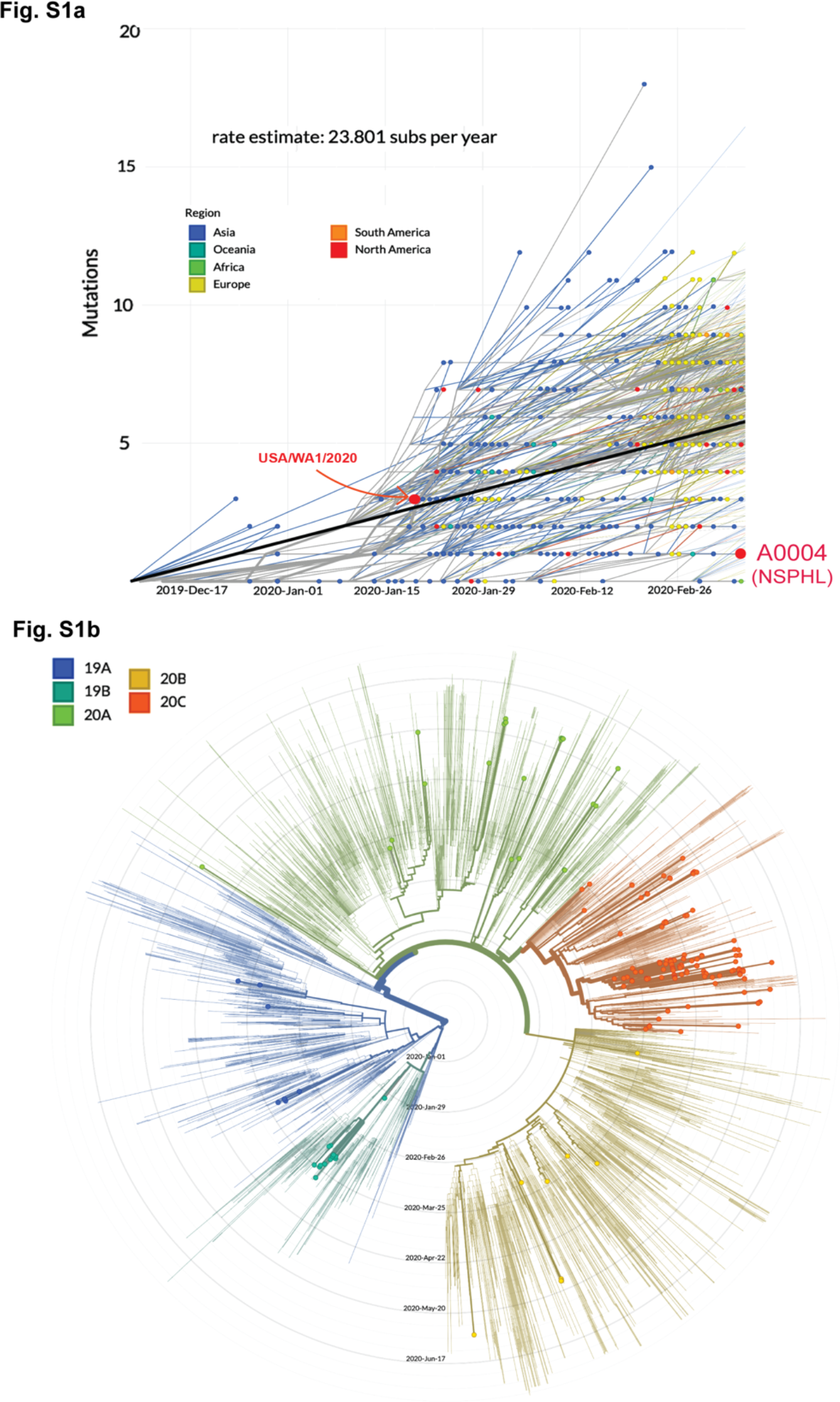
Dendrogram of Nevada specimens in context of other sequenced specimens. (a) Nucleotide mutation clock from Nextstrain.org with the SARS-CoV-2 genome isolated from Washington (USA/WA 1/2020) on January 24th and first specimen from Nevada (A0004) on March 5th indicated in red. (b) Circular dendrogram of Nevada specimens from March 6th to June 5th positioned within a subsample of global sequences from Nextstrain.org during the same time period, the larger dots indicate specimens from Nevada. The five clades are colored 19A (blue), 19B (teal), 20A (green), 20B (yellow) and 20C (orange).

**Figure S3.**
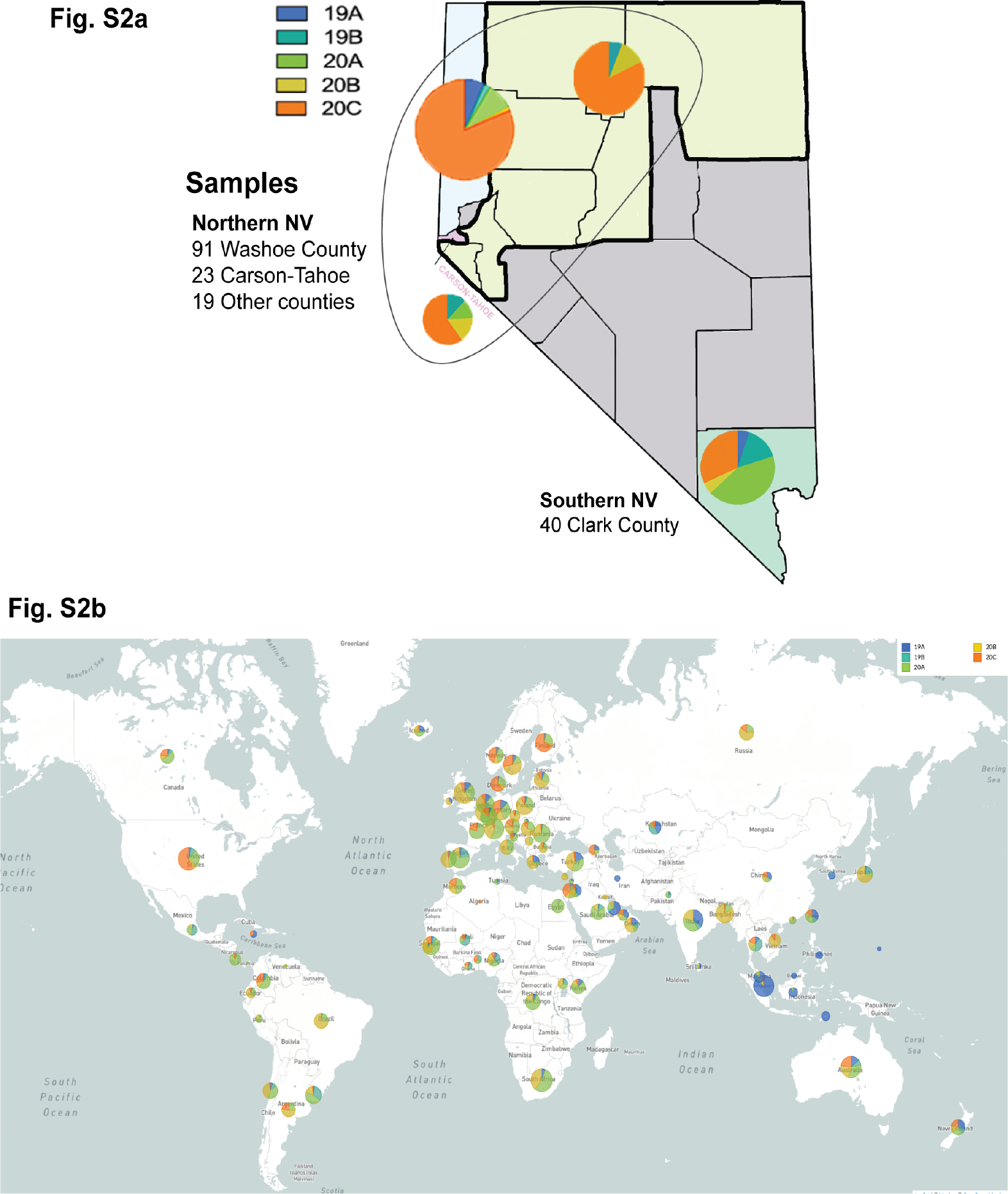
Global distribution of clades from March 6th to June 5th. Pie charts depict the proportion of the clades within (a) four main areas in Nevada, these include 91 specimens from Washoe county (upper left), 23 specimens from Carson-Tahoe (middle left), 40 specimens from Clark county (bottom right), and 19 from rural Nevada (middle). The five clades are colored 19A (blue), 19B (teal), 20A (green), 20B (yellow) and 20C (orange). (b) Pie chart of the clades in each indicated country from March 6th to June 5th generated from a subsampling of sequences deposited in Nextstrain.org. The size of the pie chart corresponds to the relative specimen number for each region.

